# Signatures of COVID-19 severity and immune response in the respiratory tract microbiome

**DOI:** 10.1101/2021.04.02.21254514

**Authors:** Carter Merenstein, Guanxiang Liang, Samantha A. Whiteside, Ana G. Cobián-Güemes, Madeline S. Merlino, Louis J. Taylor, Abigail Glascock, Kyle Bittinger, Ceylan Tanes, Jevon Graham-Wooten, Layla A. Khatib, Ayannah S. Fitzgerald, Shantan Reddy, Amy E. Baxter, Josephine R. Giles, Derek A. Oldridge, Nuala J. Meyer, E. John Wherry, John E. McGinniss, Frederic D. Bushman, Ronald G. Collman

## Abstract

**Rationale:** Viral infection of the respiratory tract can be associated with propagating effects on the airway microbiome, and microbiome dysbiosis may influence viral disease.

**Objective:** To define the respiratory tract microbiome in COVID-19 and relationship disease severity, systemic immunologic features, and outcomes.

**Methods and Measurements:** We examined 507 oropharyngeal, nasopharyngeal and endotracheal samples from 83 hospitalized COVID-19 patients, along with non-COVID patients and healthy controls. Bacterial communities were interrogated using 16S rRNA gene sequencing, commensal DNA viruses *Anelloviridae* and *Redondoviridae* were quantified by qPCR, and immune features were characterized by lymphocyte/neutrophil (L/N) ratios and deep immune profiling of peripheral blood mononuclear cells (PBMC).

**Main Results:** COVID-19 patients had upper respiratory microbiome dysbiosis, and greater change over time than critically ill patients without COVID-19. Diversity at the first time point correlated inversely with disease severity during hospitalization, and microbiome composition was associated with L/N ratios and PBMC profiles in blood. Intubated patients showed patient-specific and dynamic lung microbiome communities, with prominence of *Staphylococcus*. *Anelloviridae* and *Redondoviridae* showed more frequent colonization and higher titers in severe disease. Machine learning analysis demonstrated that integrated features of the microbiome at early sampling points had high power to discriminate ultimate level of COVID-19 severity.

**Conclusions:** The respiratory tract microbiome and commensal virome are disturbed in COVID-19, correlate with systemic immune parameters, and early microbiome features discriminate disease severity. Future studies should address clinical consequences of airway dysbiosis in COVID-19, possible use as biomarkers, and role of bacterial and viral taxa identified here in COVID-19 pathogenesis.

## Introduction

COVID-19, caused by the coronavirus SARS-CoV-2, is a global pandemic with severe morbidity and mortality, and unprecedented economic and social disruption. A striking feature of COVID-19 is the wide variance in clinical severity among infected people. Many factors correlate with COVID-19 disease severity, including age, gender, body mass index, prior comorbidities, immune responses and genetics (1–3), yet the determinants of infection outcome and pathogenic mechanisms are incompletely understood. Here we investigate the potential relationship between COVID-19 severity and the microbiome of the respiratory tract.

The respiratory tract is the site of initial SARS-CoV-2 infection and the most common site of serious clinical manifestations. Infection of any mucosal surface occurs in the context of its endogenous microbiome, and bi-directional interactions between host and microbiota commonly contribute to infection and pathogenesis. For example, influenza predisposes to secondary bacterial infection in the respiratory tract, which is responsible for much of its morbidity and mortality (4–7). Conversely, prior disruption of the normal microbiome can influence susceptibility to or pathogenesis of respiratory viruses such as influenza and respiratory syncytial virus (8–10). Few studies have addressed the respiratory tract microbiome in COVID-19 or links to outcome, though early data report evidence of dysbiosis (11, 12).

Here we investigated signatures of COVID-19 disease in the respiratory tract microbiome, analyzing 507 oropharyngeal, nasopharyngeal and endotracheal specimens from 83 hospitalized COVID-19 patients. We also collected 75 specimens from 13 critically ill patients hospitalized for other disorders. Bacterial community composition was assessed using 16S rRNA gene sequencing. Levels of commensal viruses of the human airway, specifically *Anelloviridae* and *Redondoviridae*, were quantified using qPCR. These small circular DNA viruses have been reported to vary in abundance in association with disease states and/or immune competence (13–15), so we reasoned that they might report aspects of COVID-19 disease status. Finally, we queried the relationship between the airway microbiome and immune profiles in blood. Our analysis revealed dysbiosis of the upper and lower respiratory microbiome, differences between COVID-19 and non-COVID patients, associations with systemic inflammation, and microbial signatures distinguishing COVID-19 severity.

## Materials and Methods

### Subjects

Following informed consent (IRB protocol #823392), samples were collected beginning a median of 4 days after hospitalization (generally within one week of hospitalization or identification of COVID+ status if post-admission). Oropharyngeal (OP) and nasopharyngeal (NP) swabs, and endotracheal aspirates (ETA) from intubated subjects, were obtained as previously described (16). Additional OP and NP swabs were obtained and eluted in viral transport media (VTM) for SARS-CoV-2 analysis as previously described (17). COVID-19 patients were classified clinically by maximum score reached during hospitalization using the 11-point WHO COVID-19 progression scale (18). Non-COVID subjects were patients hospitalized in the intensive care unit (ICU) with a variety of underlying disorders. Healthy controls included 30 individuals who underwent OP and NP sampling and 12 subjects who underwent bronchoscopy and bronchoalveolar lavage (BAL) previously as described (16, 19, 20).

### 16S rRNA gene sequencing and analysis

DNA extraction, 16S rRNA gene PCR amplification using V1V2 primers, and Illumina sequencing was carried out as described (21, 22). NP, OP and BAL 16S rRNA gene V1V2 sequences of healthy controls were acquired previously using the Roche 454 GS-FLX platform (16, 19, 20), which we showed yields comparable results to and can be integrated with Illumina data (16). Processing using the QIIME2 pipeline, calculations of alpha-diversity, UniFrac distances, principal coordinate analysis (PCoA), and PERMANOVA testing are detailed in Supplemental Methods. For analyses comparing groups with different numbers of samples per subject, PERMANOVA testing used specimens randomly subsampled 1000 times to one sample per patient and mean p values reported.

### Viral analysis

Extracted DNA was amplified using Phi29 DNA polymerase and random hexamers, then subject to qPCR using primers/probes that target *Redondoviridae* (RV) and *Anelloviridae* as described (15, 23). Levels of SARS-CoV-2 RNA were quantified in total RNA extracted from ETA or VTM and complete SARS-CoV-2 genome sequences were generated as recently reported (17).

### Clinical and immune data

Clinical laboratory test results were extracted from the electronic medical record. Flow cytometric cellular immune profiling of PBMC was available on a subset of subjects as described (24). The unbiased Uniform Manifold Approximation and Projection (UMAP) approach was used to distill 193 individual immune components into two principal components (24). The microbiome unweighted UniFrac PCoA was compared with blood cellular UMAP analysis using Mantel’s test and Procrustes analysis.

### Statistical analysis

Nonparametric tests were used to compare two independent groups (Wilcoxon rank-sum test), two related groups (Wilcoxon signed-rank test) and multiple groups (Kruskal–Wallis test). Spearman’s rank-order correlation was used for non-parametric correlation analysis. Fisher’s exact tests were used to test difference between two categorical variables. P values are from two-sided comparisons. P values for multiple comparisons were corrected using the Benjamini–Hochberg FDR method. Random forest classification was implemented using the randomForest package (v4.6-14) in R. Decision trees were trained on data consisting of bacterial relative abundance at the genus level and small circular DNA viruses copy numbers (*Redondoviridae* or *Anelloviridae*) in samples from the first two time points as detailed in Supplemental Methods.

## Results

### Subjects, specimens and SARS-CoV-2 analysis

The 83 COVID-19 patients (Tables 1, E1 and Figure S1) had a median age of 64 years (range 36-91) and included 39 women and 44 men. Fifty-six identified as Black (67%), 20 White (24%), 3 Asian (4%) and 4 unknown/other race (5%). All but 5 had at least one underlying major organ system comorbidity. Forty (48%) required intubation and invasive mechanical ventilation, and 20 (24%) died. Each patients’ clinical course was classified by maximal severity reached during hospitalization based on the WHO 11-point scale (18) in which hospitalized patients are level 4 or above, intubated subjects level 7 or above, and fatal outcomes are 10. Non-COVID critically ill patients (n=13) included a variety of underlying diseases, of whom 62% required intubation and 6 (46%) died. Upper respiratory tract (OP and NP) and lung (ETA) sampling was carried out serially, yielding a total of 582 specimens for microbiome analysis (507 COVID-19, 75 non-COVID). Healthy volunteers provided NP and OP (n=30) and lung (BAL; n=12) specimens (16, 19, 20).

**Table 1.** Patients studied.

|  | **COVID-19 (N=83)** | **non-COVID (N=13)** |
| --- | --- | --- |
| **Gender** |  |  |
| Female | 39 (47%) | 4 (31%) |
| Male | 44 (53%) | 9 (69%) |
| **Race / Ethnicity** |  |  |
| Black | 56 (67%) | 7 (54%) |
| White | 20 (24%) | 6 (46%) |
| Asian | 3 (4%) | 0 |
| Other/Unknown | 4 (5%) | 0 |
| Hispanic/Latinx | 0 | 1 (8%) |
| **Age** |  |  |
| Median [Min, Max] | 64 [36-91] | 60 [39-940] |
| **BMI** |  |  |
| Median [Min, Max] | 301 [17-62] | 23 [19-31] |
| **Preexisting comorbidities:** |  |  |
| Diabetes | 39 (47%) | 6 (46%) |
| Hypertension | 67 (81%) | 7 (54%) |
| Coronary artery disease | 16 (19%) | 3 (23%) |
| Stroke | 17 (20%) | 2 (15%) |
| Chronic lung disease | 34 (41%) | 5 (38%) |
| Renal disease (≥stage 4) | 15 (18%) | 3 (23%) |
| Cancer (within 6 months) | 10 (12%) | 4 (31%) |
| HIV infection | 3 (4%) | 0 |
| Organ transplant | 5 (6%) | 1 (8%) |
| Immunosuppressive therapy | 12 (14%) | 3 (23%) |
| BMI ≥35 | 27 (33%) | 0 |
| Any major comorbidity | 78 (94%) | 13 (100%) |
| **Treatment** |  |  |
| Corticosteroids | 52 (63%) |  |
| Remdesivir | 18 (22%) |  |
| Hydroxychloroquine | 39 (47%) |  |
| Convalescent Plasma | 4 (5%) |  |
| Antibacterials | 72 (87%) | 13 (100%) |
| Antifungals | 20 (24%) | 5 (38%) |
| Mechanical ventilation | 40 (48%) | 8 (62%) |
| ECMO | 5 (6%) | 1 (8%) |
| **Maximum WHO Score (examples)** |  |  |
| 4 (no supplemental O2) | 20 (24%) |  |
| 5 (low flow O2) | 8 (10%) |  |
| 6 (high flow O2) | 7 (8%) |  |
| 7 (intubated) | 2 (2%) |  |
| 8 (intubated low P/F; vasopressors) | 14 (17%) |  |
| 9 (ECMO; pressors; dialysis) | 12 (14%) |  |
| 10 (died) | 20 (24%) | 6 (46%) |

SARS-CoV-2 RNA levels were variable among subjects and sample types (Figure S2). As expected, SARS-CoV-2 RNA levels declined to undetectable over time in most patients, although several had persistently detectable RNA in ETA beyond 3 weeks post-symptom onset. There was no association between SARS-CoV-2 RNA levels and WHO score or clinical outcomes (Wilcoxon rank-sum test). Complete SARS-CoV-2 genome sequences were determined for 26 subjects as recently reported (17). All viral genomes were members of the B.1 lineage, which encodes the D614G variant in Spike and most also had the P314L variant in the RNA-dependent RNA polymerase (RdRp) located on ORF1b (25, 26).

### Respiratory tract bacterial dysbiosis in COVID-19

Bacterial communities were interrogated using primers targeting the V1V2 region of the 16S rRNA gene, which has been employed extensively for airway samples (21, 22) (Figures 1, S3). Oropharyngeal and nasopharyngeal communities of COVID-19 patients differed markedly from healthy subjects using the unweighted UniFrac metric, which compares samples based on bacterial presence-absence information (Figure 1A,B; p<0.00001 both OP and NP). We then compared COVID-19 to non-COVID patients, and COVID-19 patients to each other based on maximal severity during hospitalization. COVID-19 patients were grouped as WHO 4-6 (moderate/severe, non-intubated), WHO 7-9 (critical/intubated) and WHO 10 (fatal) (Figure 1B,C). In both oropharynx and nasopharynx, there was significant separation between groups. In pairwise comparisons, all COVID-19 groups were significantly different from the non-COVID group (FDR<0.01 OP, FDR<0.05 NP). Oropharyngeal swabs also showed separation between COVID-19 patients with moderate/severe (WHO 4-6) and critical/fatal (WHO 7-10) outcomes (FDR<0.06).

**Figure 1.**
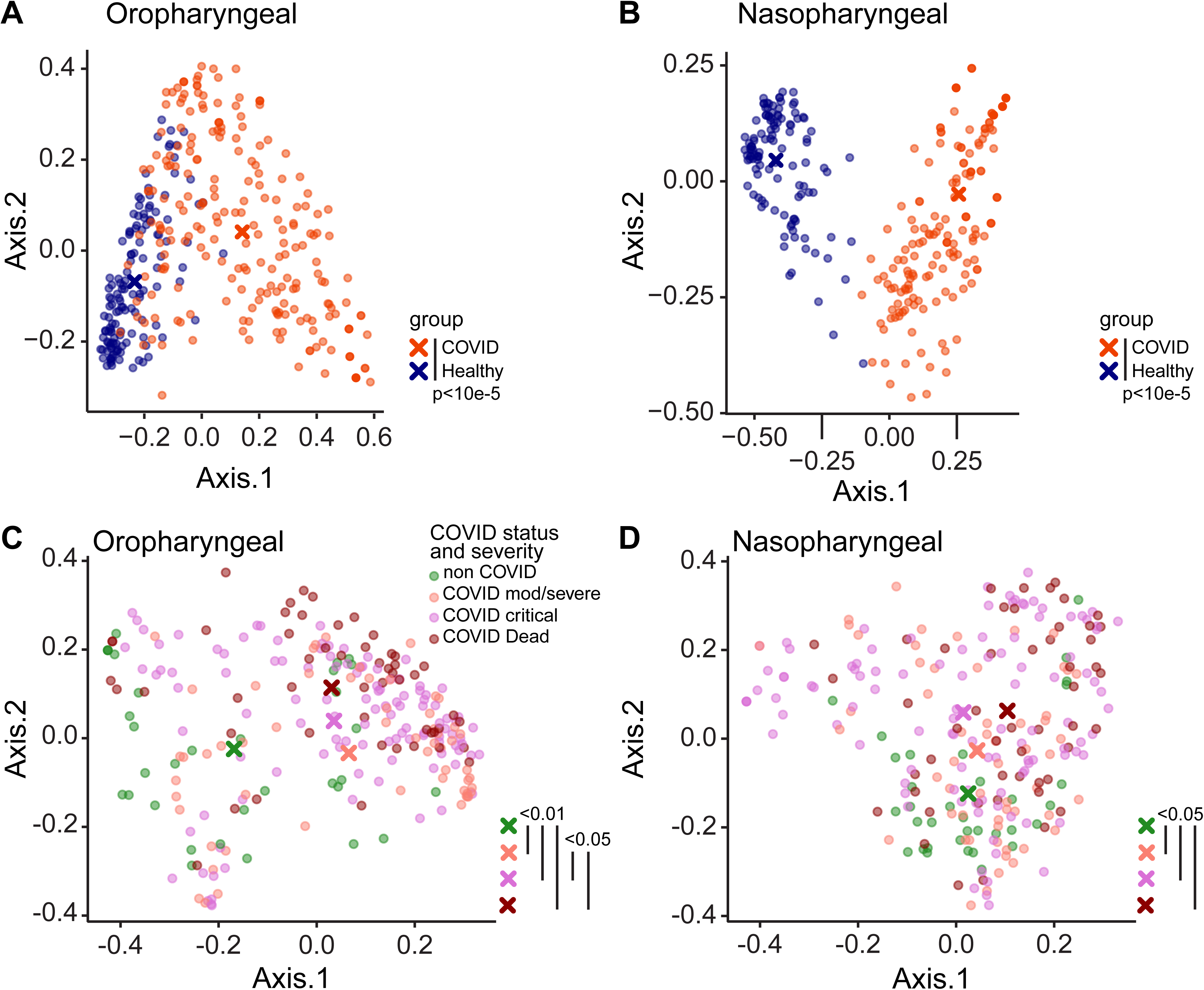
Upper respiratory tract dysbiosis in COVID-19 patients. Bacterial communities in the oropharynx (**A, C)** and nasopharynx (**B, D)** were analyzed by unweighted UniFrac. **A, B.** Communities from COVID-19 subjects compared to healthy individuals. **C, D.** Communities from hospitalized patients grouped by COVID-19 status and disease severity as moderate/severe (WHO 4-6), critical (WHO 7-9) and fatal (WHO 10). All samples are shown; P values were generated using random subsampling for each subject. The centroid for each subject group is indicated by X, and significant differences between groups indicated by bars at the right of each plot.

We compared bacterial phyla that differed between groups, using repeated random subsampling to reduce bias associated with sicker patients being hospitalized longer and having more samples (Figure 2A,B). Across groups, COVID-19 patients had lower OP abundance of Proteobacteria than non-COVID patients, and a trend toward greater abundance of Bacteroidetes (FDR=0.008 and FDR=0.06, respectively; Kruskal Wallis). NP communities showed similar trends but did not meet the FDR threshold for statistical significance. Pairwise between-group comparisons are shown in Figure 2.

**Figure 2.**
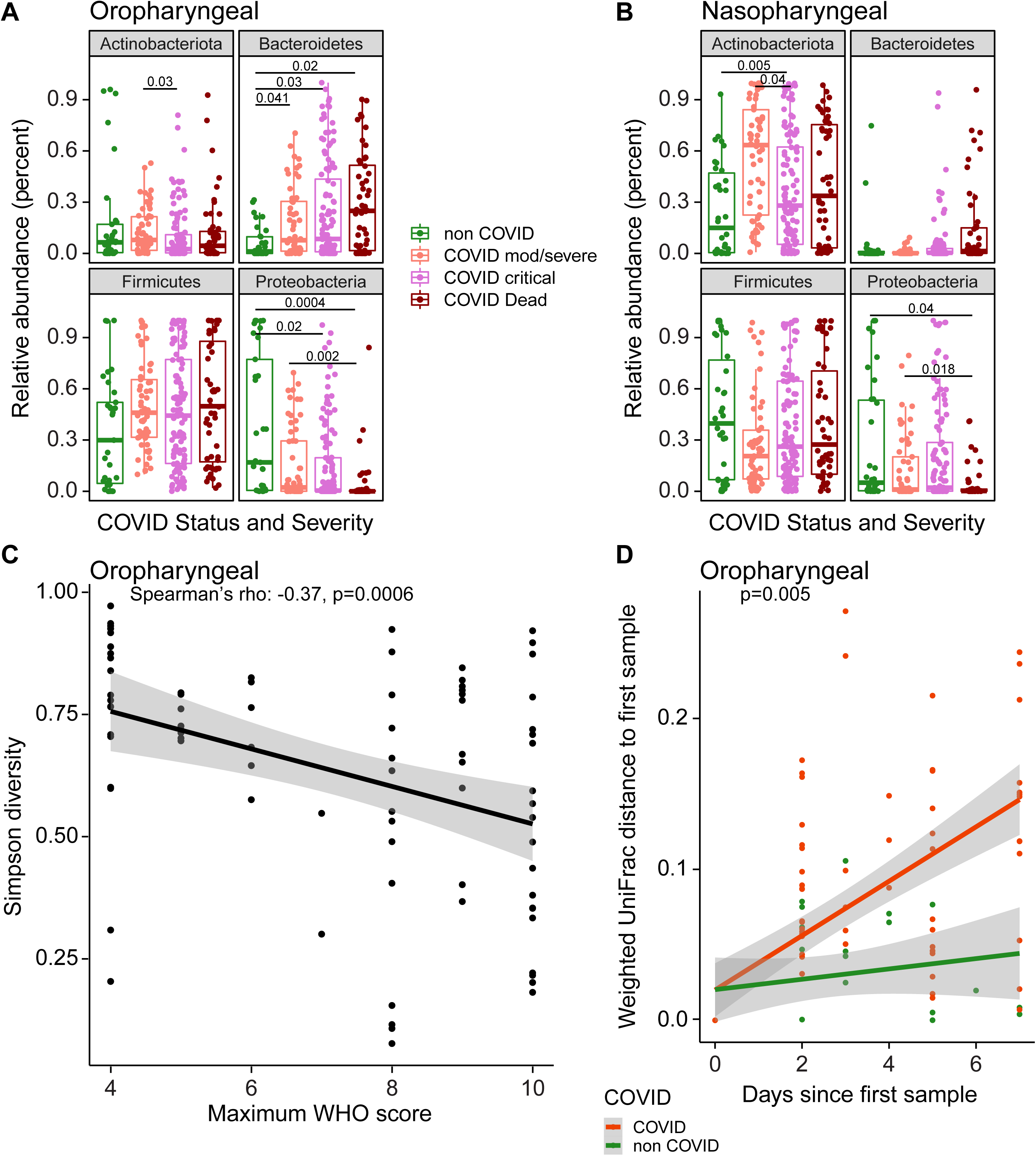
Signatures of disease severity in airway bacterial populations. **A, B.** Relative abundances of bacterial phyla, by patient disease status categories. All samples are shown; P values were generated using random subsampling for each subject and indicate Wilcoxon pairwise comparisons. **C**. Maximum WHO score reached by each patient (x-axis) versus Simpson Diversity Index in the first oropharyngeal sample obtained for each subject (y-axis). The grey shading shows the 95% confidence interval. **D**. Divergence in oral bacterial communities over time, comparing COVID-19 (red) and non-COVID (green) samples. The x-axis shows the time since the first sample, the y-axis shows the weighted UniFrac distance to the first sample. The grey shading shows the 95% confidence interval.

We then assessed the first sampling time point and found that, among of COVID-19 patients, decreased oropharyngeal Proteobacteria and Actinobacteria correlated with greater WHO score over the course of hospitalization (Spearman’s rho: −0.36 and −0.28 respectively; FDR=0.008 and 0.05 respectively). At the genus level, patients with more severe disease had significantly lower relative abundances of *Hemophilus, Actinomyces* and *Neisseria* (FDR<0.05, Figure S4), all of which are abundant in the normal oropharyngeal microbiome.

Alpha-diversity in oropharyngeal samples at the first time point also correlated with COVID-19 severity, with lower diversity associated with higher WHO score (Spearman’s rho: −0.37, p=0.0006, Figure 2C). We then assessed rate of change over time, comparing COVID-19 and non-COVID subjects. Community types were summarized using weighted UniFrac values, which scores bacterial abundances, and divergence over time from the subject’s initial sample was calculated (Figure 2E). The rate of change in oropharyngeal bacterial community structure was significantly greater in COVID-19 than non-COVID subjects (p=0.005, Kruskal-Wallis), indicating that COVID-19 patients experience greater destabilization of bacterial communities during their illness. Significant differences were not seen with unweighted UniFrac, emphasizing that differences were primarily associated with changes in community proportions rather than membership.

### The lung microbiome in intubated COVID-19 patients

The lung microbiome was interrogated in endotracheal aspirate (ETA) samples from 24 intubated subjects (Figure 3). ETA from COVID-19 patients had markedly lower diversity than lung communities from healthy people (Simpson index 0.56 vs 0.86; p=5.1×10^−6^, Wilcoxon rank-sum test, Figure 3A). Lineages were heterogeneous and revealed both common respiratory pathogens (*Staphylococcus*, *Klebsiella*, *Stenotrophomonas*) and taxa typical of upper respiratory tract communities (*Corynebacterium*, *Prevotella*).

**Figure 3.**
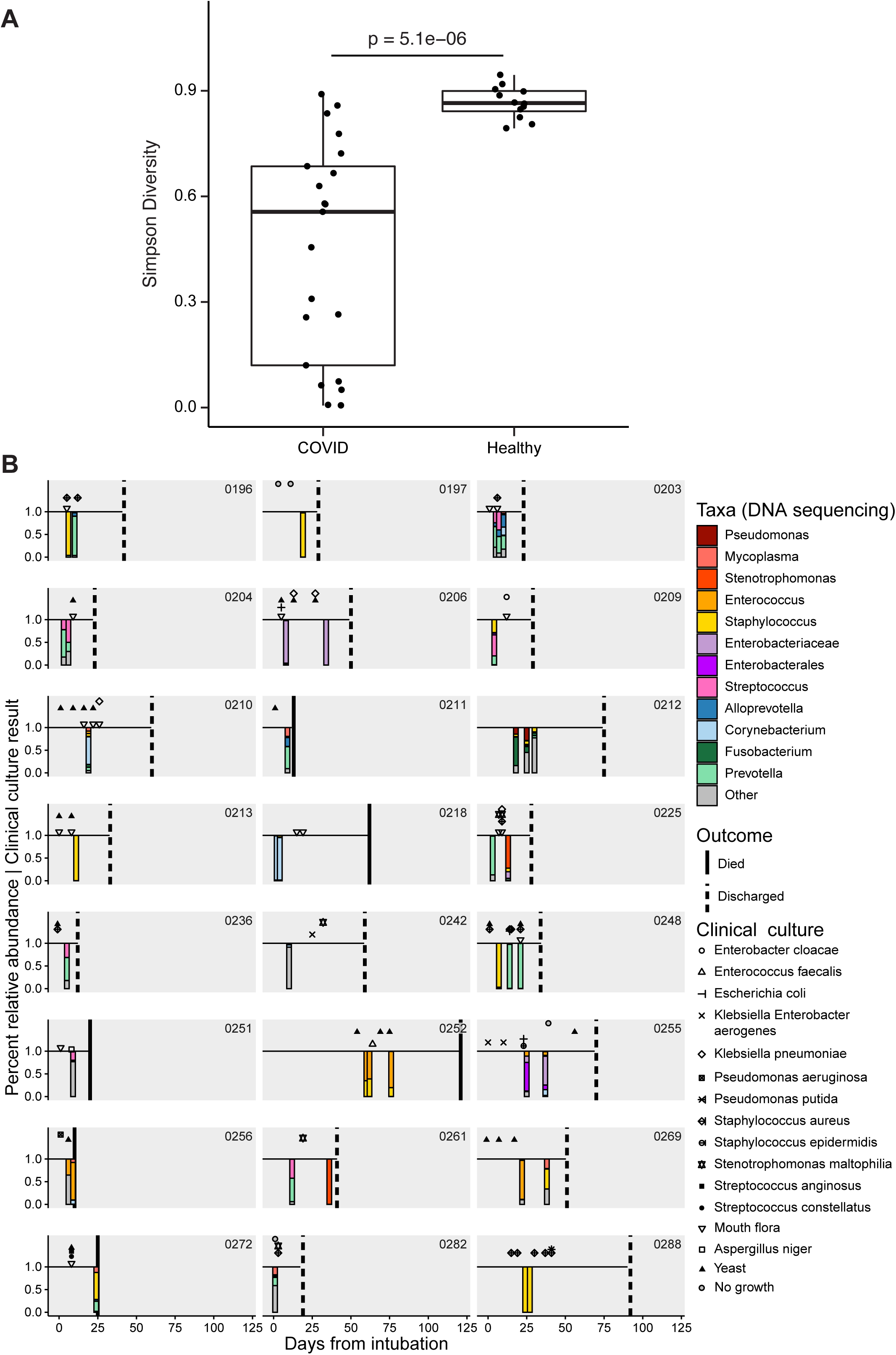
The lower respiratory tract microbiome in intubated COVID-19 patients. **A.** Simpson diversity of ETA samples from COVID-19 patients and healthy subjects’ BAL. For COVID-19 patients with multiple samples only the first ETA sample was used. **B.** Timeline of subjects and samples, with results of endotracheal aspirate 16S sequence analysis shown below the line as stacked bar plots (color key to the right), and clinical culture results shown above the line (key to symbols at right). Taxa are indicated at the lowest taxonomic level assigned by the QIIME/SILVA pipeline, but further identification by BLAST alignment revealed unassigned *Enterobacteriaceae* to be *Klebsiella aerogens* and *Enterobacterales* to be *Eschericia coli*.

Six of 24 subjects had one or more ETA samples dominated by *Staphylococcus* (subjects 196, 197, 213, 248, 272, 288), and another three revealed *Staphylococcus* as a prominent minority constituent (209, 252, 269). Of the 5 subjects with *Staphylococcus* domination who had respiratory culture within 1 week of sampling, only 3 had *S. aureus* identified by culture, suggesting that either 16S sequencing is more sensitive than culture, or dominant *Staphylococcus* is not *S. aureus*. Three subjects had ETA samples dominated by *Enterococcus* (252, 256, 269). Other respiratory pathogen taxa that dominated smaller numbers of samples (two patients each) included *Stenotrophomonas*, *Enterobacteriaceae* (identified as *Klebsiella aerogens* by BLAST) and *Enterobacterales* (*Eschericia coli* by BLAST).

Among subjects who had serial ETA samples, several showed stable composition over time (203, 204,206, 218, 252, 288), while some demonstrated modest or gradual compositional evolution (212, 255, 256). In contrast, several showed marked changes between longitudinal samples (196, 225, 248, 261, 269). Thus, the lower respiratory tract microbiome in critically ill intubated COVID-19 patients is low diversity, can be dominated by either pathogens or normal upper respiratory taxa, may have a predilection for *Staphylococcus*, and can be highly dynamic.

### Commensal DNA viruses are associated with disease severity

We next assessed the presence of two airway commensal DNA viruses, *Anelloviridae* and the recently-described *Redondoviridae* (13–15). Figure 4A shows longitudinal data for *Anelloviridae* and *Redondoviridae* combined with bacterial taxa. At early time points, both *Anelloviridae* and *Redondoviridae* in oropharyngeal samples was positively associated with intubation during hospitalization (Figure 4B,C; Table E4; FDR=0.02 for both, Fischer’s exact test with FDR correction). The commensal DNA viruses were also associated with higher WHO score (Table E4).

**Figure 4.**
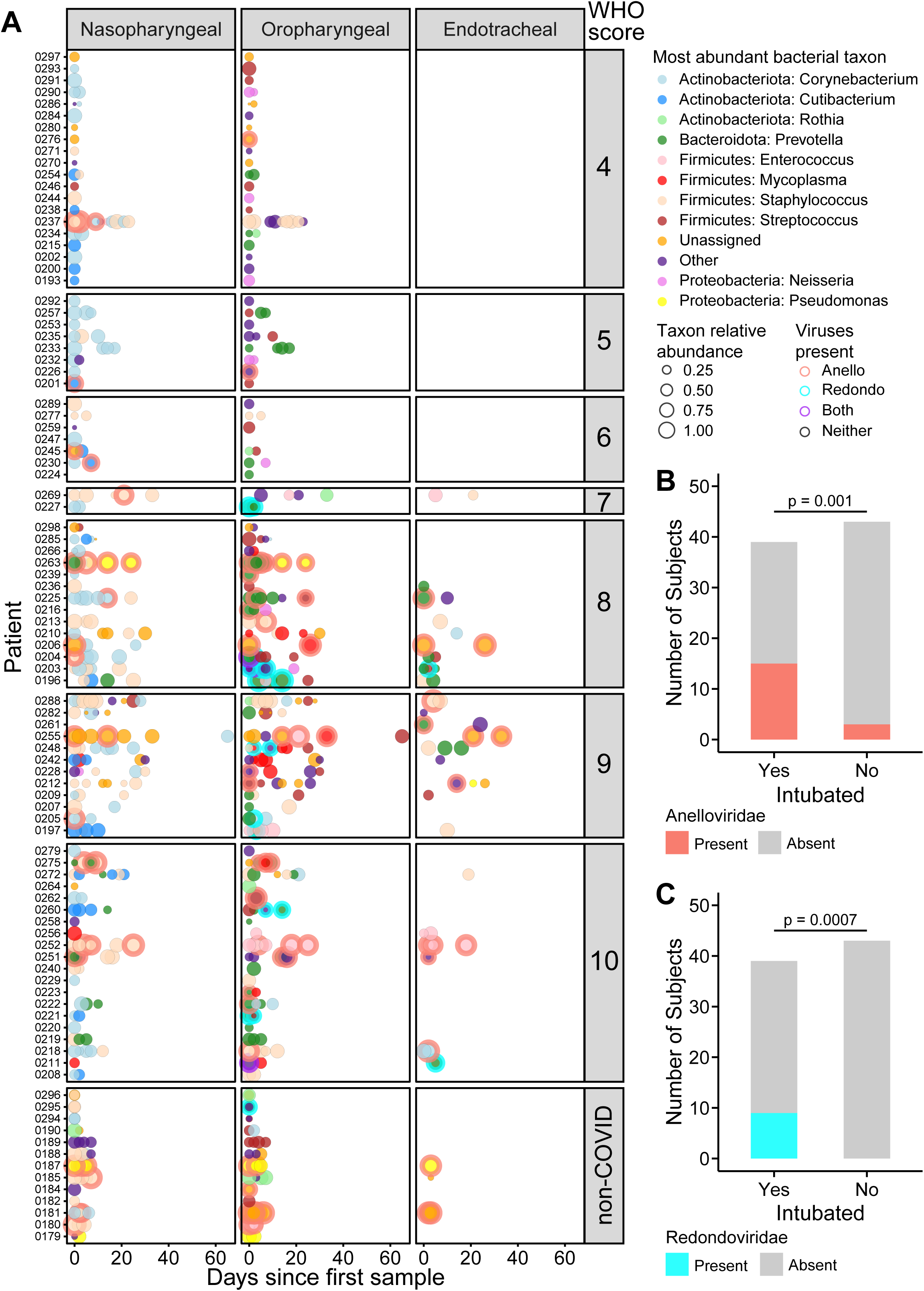
Bacterial dominance and commensal viruses in airway microbial communities. **A**. Summary of the most abundant taxa in each subject at different time points in nasopharyngeal, oropharyngeal and endotracheal communities. The x-axis shows days since first sample; each row shows a different patient. Subjects are grouped based on COVID-19 WHO score, with non-COVID patients at the bottom. The types of bacteria are indicated by the color code to the right and the size of the circle indicates the relative abundance of that dominant bacterial taxon. Detection of *Anelloviridae* and/or *Redondoviridae* is indicated by the ring around some disks and color coded as indicated to the right. **B, C**. Detection of *Anelloviridae* and *Redondoviridae* in intubated versus non-intubated patients. To control for longer sampling period in sicker patients, detection in only the first two time point samples were considered.

### The respiratory microbiome is related to systemic immune responses

We asked whether airway microbiome communities were related to systemic immune or inflammatory features. The ratio of lymphocytes and neutrophils has been associated with COVID-19 severity and outcomes (27, 28). We found that lower lymphocyte-to-neutrophil ratio (LNR) was associated with both lower diversity (FDR=0.03, r=0.23, Spearman correlation; Figure 5A) and composition of the oropharyngeal microbiome (UniFrac second principal coordinate: FDR=0.01, r=0.32 weighted; FDR=0.008, r=0.35, unweighted; Spearman correlation; Table E5). As expected, LNR correlated inversely with disease severity (FDR=3.5×10^−5^, r=-0.6, Spearman correlation; Figure 5B).

**Figure 5.**
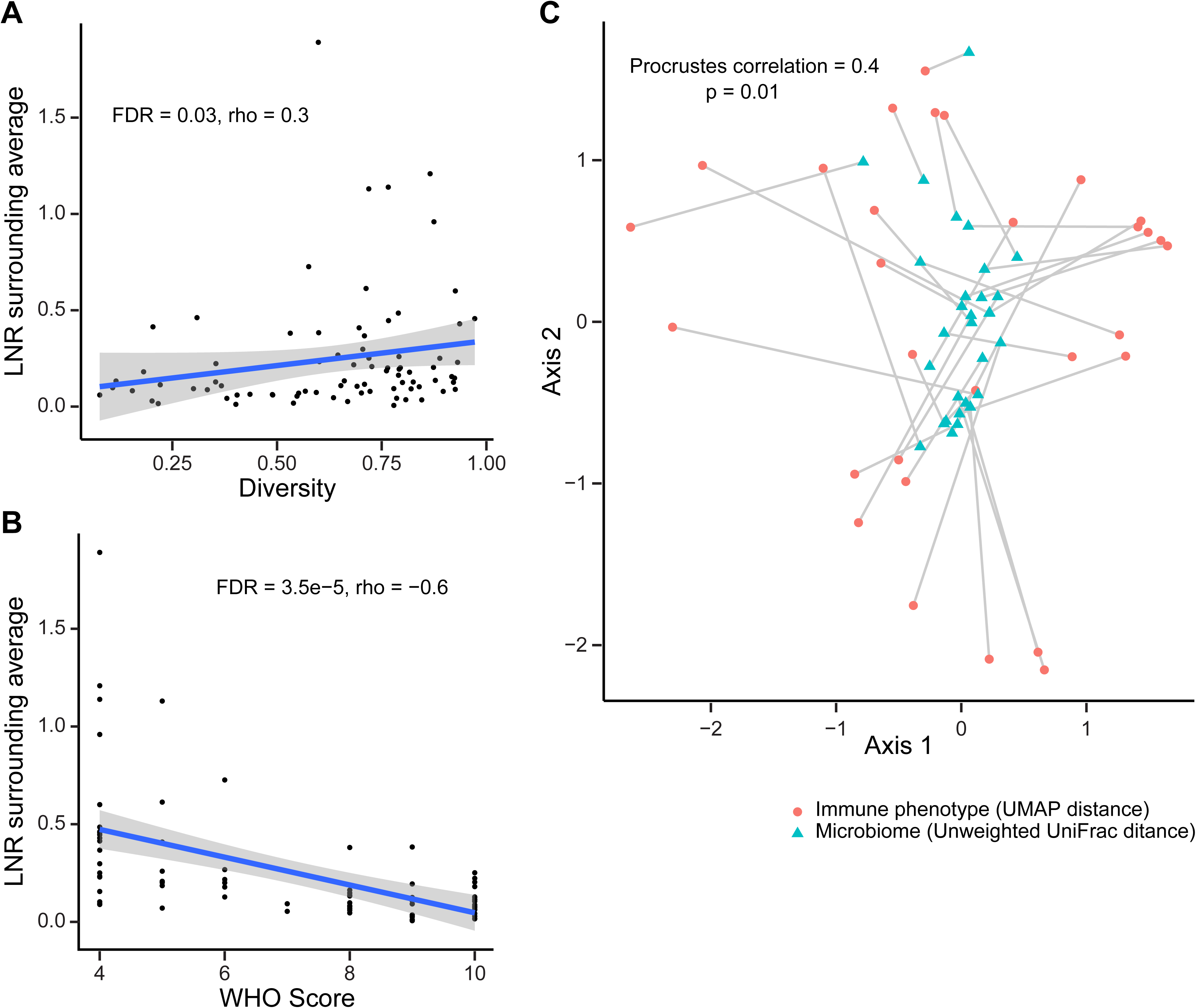
Relationship between oropharyngeal microbiome communities and systemic immune features. **A**. Oropharyngeal microbiome diversity at the first time point sampled is plotted against the blood lymphocyte/neutrophil ratio at the time of sampling. **B**. Blood lymphocyte/neutrophil ratio at time of oropharyngeal sampling (from panel A) is plotted against maximum WHO score during hospitalization. **C**. Procrustes analysis in which the UMAP immune profile plot and unweighted UniFrac microbiome plot are overlaid. The immune and microbiome profiles from individual subjects are connected by a line

We then investigated peripheral blood mononuclear cell (PBMC) phenotyping that was available on 34 of the subjects (Table E1) co-enrolled in a deep immune profiling study of COVID-19 patients (24). That study assessed 193 individual cellular immune features and integrated them in a high-dimensional immune phenotype analysis (Uniform Manifold Approximation and Projection; UMAP) that reduced the immune features to a two-dimensional landscape and created compacted meta-features reflected in the two components.

We initially tested a limited set of individual B cell and T cell markers and found no association with the bacterial or viral microbiome that reached significance after FDR correction (Table E7). To encompass the multiple immune parameters, we compared global immune patterns to overall microbiome profiles by comparing the UMAP distance matrix generated by the 193 immune components to the respiratory microbiome unweighted UniFrac distance matrix. This revealed a significant correlation between the OP microbiome distance matrix and the systemic immune profile distance matrix (p<0.001; Mantel’s test). We also applied a Procrustes analysis, which provides visualization of the overlay of the two multidimensional matrices (Figure 5C), and which also revealed a significant correlation (Procrustes correlation 0.4; p=0.01). Thus, the oropharyngeal microbiome composition is globally correlated to systemic immune cell composition.

### Machine learning identifies signatures associated with COVID-19 severity

Lastly, we sought to identify microbiome features most associated with disease severity, employing the random forest machine learning algorithm. This analysis incorporated abundances of bacterial taxa, bacterial community features and commensal DNA viruses in OP or NP samples to discriminate patients who needed intubation and WHO score (Figure 6). We used the first two samples for each patient to allow more homogenous comparison between patients sampled for different durations.

**Figure 6.**
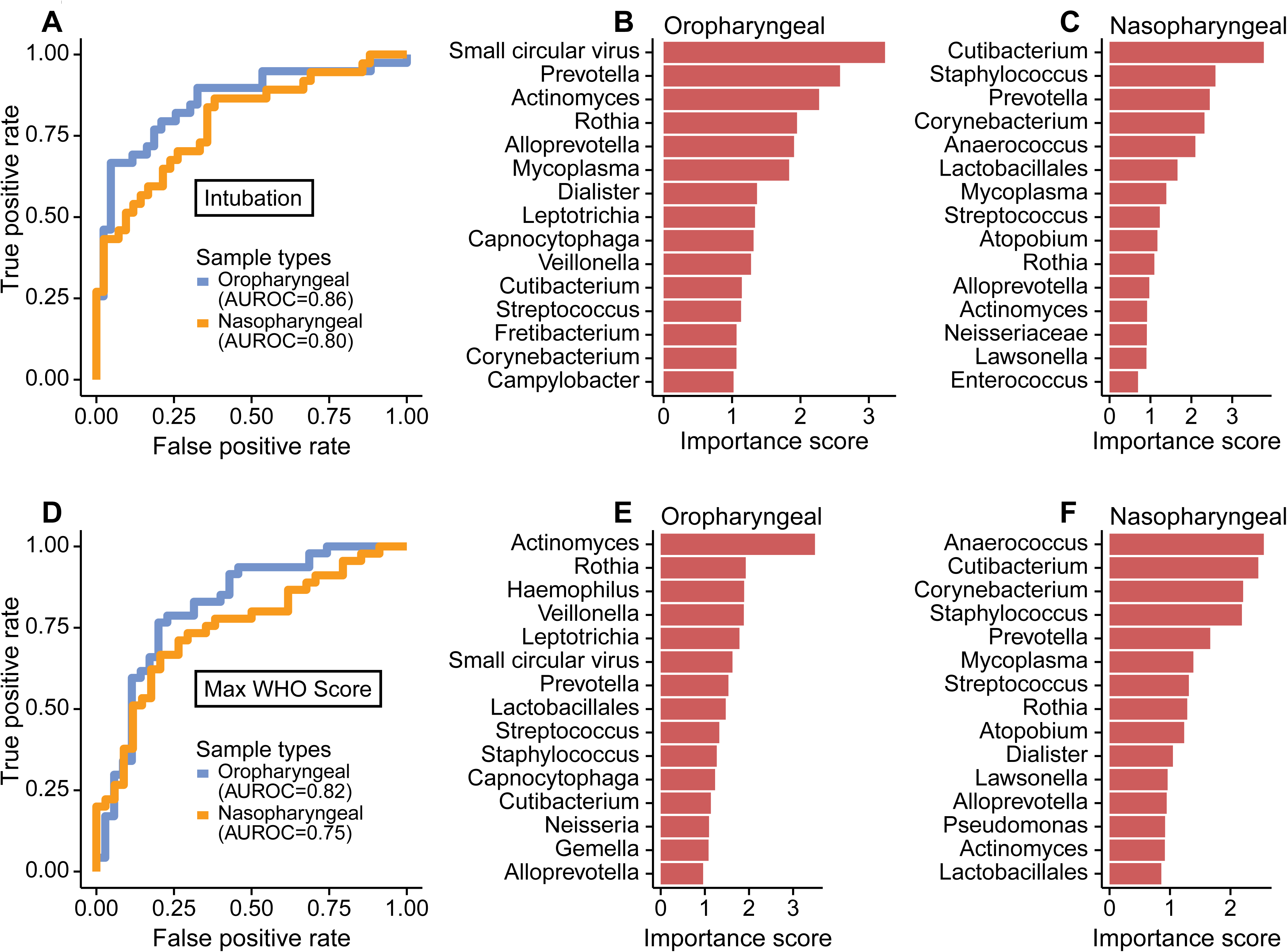
Random forest to detect signatures of severity in the SARS-CoV-2-infected subjects. **A**. Receiver operating characteristic (ROC) curve of random forest classification on patient intubation status using OP and NP samples. Top 15 most important predictors in classifying patient’s intubation status using OP (**B**) and NP (**C**) samples are shown. **D**. ROC curve of random forest classification on disease severity using both OP and NP samples. Top 15 most important predictors in classifying disease severity using OP (**E**) and NP (**F**) samples are shown.

Both NP and OP microbiome data discriminated between patients requiring intubation or not (Area Under the Receiver Operating Characteristics (AUROC)=0.80 and 0.86, respectively; Figure 6A). The feature that contributed most to clinical status discrimination was the presence of small circular DNA viruses for OP samples, with higher levels of the viruses associated with intubation (Figure 6B). Also positively associated with intubation were *Prevotella* and *Mycoplasma*. Lower random forest accuracy rates were observed in NP compared to OP samples, but more than 50% of the most important taxa were in common between the two sample types (Figure 6C). OP and NP data also discriminated disease severity (WHO score 4-6 vs 7-10, AUROC=0.82 and 0.75, respectively, Figure 6D). Bacterial lineages rather than the DNA viruses showed the strongest discriminatory power for WHO score (Figure 6E,F). Thus, these data define signatures of microbial activity associated with intubation and COVID-19 severity.

## Discussion

We found marked disruption of the oropharyngeal microbiome in hospitalized COVID-19 patients, which differed from non-COVID patients and was associated with disease severity. COVID-19 patients also showed greater destabilization of the microbiome over time than non-COVID patients. Endotracheal samples in intubated COVID-19 patients were low diversity and revealed frequent outgrowth of potential respiratory pathogens, particularly *Staphylococcus*. Oropharyngeal microbiome communities were associated with blood leukocyte populations and global PBMC immune profiles. Small DNA commensal viruses of the respiratory tract also differed among COVID-19 patients based on whether or not intubation was required. Together, the combination of bacterial and viral features at early time points had high classifier accuracy in distinguishing intubated versus non-intubated patients and clinical status reached over the course of hospitalization.

Several possibilities could account for the association between the oropharyngeal microbiome and COVID-19 severity. Dysbiosis of the vaginal microbiome and consequent inflammation is strongly associated with sexual acquisition of HIV infection (29). In the respiratory tract, the local microbiome appears to regulate mucosal immune tone (30), and local respiratory tract inflammation affects susceptibility to RSV infection and disease severity (9, 10). ACE2, the receptor for SARS-CoV-2 is an interferon-stimulated gene (31), and thus could be modulated by respiratory microbiome. While we did not have measures of local lung inflammation or immunity, we found that cellular immune profiles in blood correlated with oropharyngeal microbial communities in patients for whom both were available. Furthermore, the lung microbiome derives largely from that in the oropharynx (16) and could also be affected by OP microbiome profiles. It would be useful for future studies to investigate whether distinct respiratory microbiome profiles play a role in regulating SARS-CoV-2 infection or host response, including the propensity for infection to propagate from the upper to lower respiratory tract.

Other mechanisms could potentially link the microbiome and COVID-19 disease. Commensal bacteria with heparinase activity are reported to alter SARS-CoV-2 binding to target cells (32), but we found no differences in predicted heparinase activity based on imputed bacterial metagenomes between COVID-19 and non-COVID patients or those with different disease severity (data not shown). Conversely, SARS-CoV-2 infection might itself alter the local microbiome through inflammation or other mechanisms. It will be important to distinguish these pathways, and determine whether interventions to modify the microbiome could prevent infection or diminish disease severity.

Our longitudinal analysis revealed greater destabilization of the oropharyngeal bacterial microbiome in COVID-19 than non-COVID patients. This finding is a particularly striking given that non-COVID patients were overall sicker (all in the ICU; 40% mortality) than COVID-19 patients (both ICU and non-ICU patients; 24% mortality). While use of antibacterial drugs could differentially affect the microbiome, both COVID-19 and non-COVID patients received extensive antibiotic treatment (Fig. S1). Other interventions that might impact the microbiome, such as intubation, were also not more frequent in COVID-19 patients. This observation is consistent with the possibility that SARS-CoV-2 infection of the respiratory mucosa might itself also drive changes in microbiome communities.

The small commensal DNA viruses, *Anelloviridae* and *Redondoviridae*, were the top-ranking microbiome features in OP samples for distinguishing patients who required intubation. Prevalence in human adult populations ranges from 67-100% for *Anelloviridae*, which are found in blood and many tissues including the respiratory tract. *Anelloviridae* levels are typically elevated in immunocompromised states (13, 14). *Redondoviridae* are a recently-described family of viruses that appear restricted to the human oral and respiratory tract and have a prevalence of up to 15% (15, 33). *Redondoviridae* levels are elevated in the airway of intubated patients and in oral samples of periodontitis patients (15, 33), suggesting that they may be barometers for disease activity in some conditions. While small circular DNA viruses were the most powerful discriminators of intubation (Fig. 6B), they were not as strong as bacterial composition in predicting overall clinical severity (Fig. 6E). Since some subjects reached higher WHO scoring without intubation (e.g., due to vasopressor use and/or renal failure), this result suggests that these viruses may be specifically associated with intubation.

Bacterial superinfection in COVID-19 is an area of great interest and importance (34–36). Our lung microbiome analysis revealed complex and patient-specific patterns that were often dynamic. High relative abundance was seen for several anticipated pathogens, notably *Staphylococcus* (9/24 subjects). This molecular profiling of the lung microbiome is concordant with culture-based studies highlighting *Staphylococcus aureus* as an emerging co-pathogen in COVID-19 (37–39). Bacterial superinfection with *Staphylococcus* is a long-recognized consequence of influenza and important cause of morbidity and mortality (6, 7), so further studies will be useful to determine if similar mechanisms operate during COVID-19. We also noted several patients with high relative abundances of *Enterococcus*, which was recently described as a common cause of bloodstream infection in critically ill COVID-19 patients (40).

Microbiome communities were associated with systemic immune profiles. The ratio of lymphocytes to neutrophils is a marker of COVID-19 severity and outcomes (27, 28), and correlated with oropharyngeal microbiome diversity and composition. The overall structure of the oropharyngeal microbiome was also correlated with global cellular immune profiles in subjects for whom deep cellular immune profiling was available. These immune profiles are also associated with COVID-19 severity (24). It is unclear whether the respiratory tract microbiome directs systemic inflammatory responses, inflammatory response shapes the microbiome, or whether both are responsive to other factors associated with disease severity.

Our study has several limitations. Many patients presented quite ill, and when initially sampled were already intubated or had reached high WHO scores. Thus, the ability to distinguish clinical course based on microbiome markers may not necessarily reflect prior predictive power. Our patients were heterogenous, with extensive use of antibiotics that could influence the bacterial microbiome. We enrolled subjects early in the COVID-19 pandemic when clinical management and outcomes may not have been optimal. We did not analyze local lung mucosal immune/inflammatory markers, and systemic immune profiling was available for only a subset of patients. There is no gold standard for diagnosis of bacterial pneumonia superinfection in this population, limiting the ability to definitively link ETA findings. Finally, lower respiratory tract microbiome information was only available from the patients who were intubated.

In summary, we report profound dysbiosis of the respiratory tract bacterial and viral microbiome in hospitalized COVID-19 patients, which differs from that of non-COVID patients, exhibits accelerated destabilization over time, and associates with disease severity and systemic immune profiles. In intubated patients the lung microbiome is dysbiotic with frequent enrichment of *Staphylococcus*. The small commensal viruses, *Anelloviridae* and *Redondoviridae*, were the strongest discriminators of patient intubation. This work provides a basis for further studies to delineate mechanisms linking the respiratory tract microbiome and outcomes, and provides potential biomarkers to assess and/or predict clinical course.

## Supporting information

All Supplemental Tables

## Data Availability

Full data availability will be made upon final publication, with relevant data obtainable from NCBI sequence read archive and from online supplementary files.

## Acknowledgements

We are grateful to patients who provided specimens, R. Hermann and R. Trotta and the Penn Medicine nursing staff for assistance with sample collections, S. Lynch and the Penn Data Store, members of the Bushman and Collman laboratories for valuable input and suggestions, and L. Zimmerman for help with artwork and manuscript preparation.

## ONLINE DATA SUPPLEMENT

### Supplementary Methods

#### Human subjects

Following informed consent obtained under protocol #823392 approved by the University of Pennsylvania IRB, samples were collected at the Hospital of the University of Pennsylvania beginning on March 23, 2020 (two weeks after the first case was reported in Philadelphia), continuing through the first wave of the epidemic, and ending on July 10, 2020. Most patients were hospitalized for COVID-19 but a few were admitted for other reasons and found to have SARS-CoV-2 infection after hospitalization. Collections began a median of 4 days following hospitalization (generally within one week of hospitalization or identification of COVID+ status if post-admission) and continued 2-3 times weekly until discharge, death, or change in status precluding respiratory tract collections (e.g., noninvasive ventilation modalities) or 30 days from enrollment. Oropharyngeal (OP) and nasopharyngeal (NP) samples were obtained using flocked swabs (Copan Diagnostics) and endotracheal aspirate (ETA) samples were obtained from intubated patients by suction as previously described (1). Swabs and ETA were frozen (−80°C) within 1 hour of collection and stored until extraction. For some collection days (typically during the first 1-2 weeks of enrollment), additional OP and NP swabs were obtained and eluted in viral transport media (VTM) for SARS-CoV-2 analysis as previously described (2). COVID-19 patients were classified clinically based on the maximum score reached during hospitalization using the 11-point ordinal WHO COVID-19 progression scale (3). Non-COVID control subjects, who were all hospitalized in the intensive care unit (ICU) with a variety of underlying disorders, were consecutive consenting patients admitted to the ICU in two periods (September/October 2019 and July 2020) (Table E1).

Total patient and sample numbers subjected to 16S rRNA gene sequencing were: COVID-19 patients: 83; samples: 507 (OP: 226; NP: 221; ETA: 60); Non-COVID patients: 13; samples: 75 (OP: 34; NP: 34; ETA: 7); Sequencing control samples: 94 (negative controls plus synthetic positive controls (4)) (Table ES2).

#### 16S rRNA marker gene sequencing and analysis

DNA extraction, PCR amplification and sequencing were carried out as previously described (5, 6), with some modifications. DNA was extracted from swabs or 200 μL ETA using DNeasy PowerSoil or PowerSoil Pro kits (Qiagen), incorporating a 95°C x 10 minutes incubation to inactivate SARS-CoV-2. Sequencing libraries were prepared using Q5 Polymerase (New England Biolabs) and primers targeting the V1V2 regions of the 16S rRNA gene (27F and 338R; Table E8). The resulting libraries were quantified using the Quant-iT PicoGreen Assay Kit (Invitrogen), pooled in equimolar quantities, and then sequenced with 250-bp paired-end reads using the 500 cycle Rapid v2 SBS and PE Cluster HiSeq kits (Illumina) on a HiSeq 2500 in rapid run mode.

16S rRNA gene V1V2 region sequencing data were analyzed using the QIIME2 pipeline as described (7). Demultiplexed sequencing reads were imported into QIIME2 pipeline, and DADA2 was used for sequence quality filtering and denoising to generate a feature table (8). Samples with less than 1000 assigned reads were excluded from further analysis. Taxonomy was classified using the naïve Bayes classifier from QIIME2 and the SILVA database (v138.1) (9).

Alpha diversity and UniFrac distances were calculated using the vegan (v2.5-6) package in R (v4.0.2). The principal coordinate analysis (PCoA) was performed using the ape package (v5.3) in R with UniFrac distances. The vegan package was used for PERMANOVA tests. Because patients with more severe COVID-19 had a larger average number of samples than those with more moderate disease, patients were randomly subsampled to one sample per patient 1000 times when calculating PERMANOVA tests, and mean p values were reported.

To determine the rate at which sample composition diverged within a patient, we compared weighted UniFrac distance to the first sample from each patient in each sample type. To account for different sampling duration, only samples from the first 7 days were included. A linear slope was calculated for each patient, and these slopes were compared using a Kruskal-Wallis non-parametric test, so that patients with a larger number of samples were not more heavily weighted.

16S rRNA gene V1V2 region sequencing data from NP, OP and lungs (bronchoalveolar lavage; BAL) of healthy controls were previously reported (1, 10, 11). These data were acquired using the Roche 454 GS-FLX platform; our group previously showed that differences in sequencing results between the Illumina and Roche systems are minimal for these samples (1). Healthy NP, OP and BAL 16S rRNA gene reads were assigned taxonomy directly, without preprocessing, using the naïve Bayes classifier from QIIME2. When calculating unweighted UniFrac distances between these samples and samples from our current study, a threshold of 1% abundance was used to remove rare taxa from our data, to account for differences in sequencing depth between the two datasets.

#### qPCR to detect small circular DNA viruses

Total microbial DNA was amplified using Phi29 DNA polymerase (New England BioLabs) and random hexamers with the following program: 35°C for 5 minutes, 34°C for 10 minutes, 33°C for 15 minutes, 32°C for 20 minutes, 31°C for 30 minutes, 30°C for 16 hours and 65 °C for 15 minutes. Each 20 μL reaction contained 10 units Phi29 DNA polymerase, 0.1 mg/mL BSA, 1X Phi29 buffer, 2 uM random hexamers, 1 mM dNTP, and 1 μL of DNA. QPCR was performed using TaqMan Fast Universal PCR (Thermo Fisher Scientific) on a QuantStudio 3 Real Time PCR System (Applied Biosystems) with the following program: 20 sec at 95°C for 1 cycle, and 40 cycles of 95°C for 3 sec and 60°C for 30 sec. Each reaction contained 900 nM of each primer and 250 nM probe. Primers and probes that target *Anelloviridae* type species Torque Teno Virus (TTV) and *Redondoviridae* (RV) have been described previously (12, 13). Sequences are listed in Supplementary Table E8. qPCR replicates were performed in triplicate and the average genome copy number was used. Mean values are in Table E3.

#### qPCR to quantify levels of SARS-CoV-2 RNA

Levels of SARS-CoV-2 RNA were quantified as described (2), RNA was extracted from 140 μL of swab eluate, neat ETA or saliva using the Qiagen QIAamp Viral RNA Mini Kit. The RT-qPCR assay used the CDC 2019-nCoV_N1 primer-probe set (2) and sequences are listed in Table E8. RT-qPCR reactions were prepared as follows: 8.5 μl dH_2_O, 0.5 μl N1-F (20 μM), 0.5 μl N1-R (20 μM), 0.5 μl N1-P (5 μM), 5.0 μl TaqMan™ Fast Virus 1-Step Master Mix were combined per reaction. 5 μl of extracted RNA was added to 15 μl of prepared master mix for a final volume of 20 μl per reaction. Final concentrations of 2019-nCoV_N1-F and 2019-nCoV_N1-R primers were 500nM and the final concentration of the 2019-nCoV_N1-P probe was 125nM. The assay was carried out using an Applied Biosystems™ QuantStudio™ 5 Real-Time PCR System. The thermocycler conditions were: 5 minutes at 50°C, 20 seconds at 95°C, and 40 cycles of 3 seconds at 95°C and 30 seconds at 60°C.

#### Clinical and immune data

Results from clinical laboratory tests performed during the patients’ hospitalization were extracted from the electronic medical record. For lymphocyte and neutrophil values, which are measured frequently, the average value of the three days surrounding the date of microbiome sampling was used. Cellular immune profiling data was acquired on peripheral blood mononuclear cells using flow cytometry as described (14). Cell subsets queried for associations with microbiome variables are listed in Table E7. The unbiased Uniform Manifold Approximation and Projection (UMAP) approach was used to distill 193 individual immune components into two principal components (14). The microbiome unweighted UniFrac PCoA was compared with blood cellular UMAP analysis using Mantel’s test. Procrustes analysis was performed using the vegan package (v2.5-5) in R.

#### Statistical analysis

Statistical tests were conducted using R (v4.0.2). Nonparametric tests were used to compare two independent groups (Wilcoxon rank-sum test), two related groups (Wilcoxon signed-rank test) and multiple groups (Kruskal–Wallis test). A Spearman’s rank-order correlation was used to carry out non-parametric correlation analysis. Fisher’s exact tests were used to test the difference between two categorical variables. P values are from two-sided comparisons. P values for multiple comparisons were corrected using the Benjamini–Hochberg FDR method. P<0.05 or FDR-corrected P<0.05 was considered significant. All acquired data were included in analyses. Figures were generated using the R packages ggplot2 (v3.3.2).

#### Random forests

Random forest classification was implemented using the randomForest package (v4.6-14) in R. The decision trees were trained on the data consisting of bacterial relative abundance at the genus level (genera with abundances greater than 10% in at least one sample were selected) and small circular DNA viruses copy numbers (*Redondoviridae* or *Anelloviridae*). The samples from the first two time points were interrogated to control for greater sampling duration of sicker subjects; the sample with the highest commensal DNA virus level was selected from each pair. Binary variables, such as intubated or not intubated, were analyzed using classificatory random forest classification. Discriminating predictors were identified by random forest using importance values, which were calculated as mean decrease in Gini index for classification random forests. Bootstrapped iterations were performed to obtain an estimate of the misclassification rate. Receiver operating characteristic (ROC) curves, which plot the true positive rate versus false positive rate for all possible threshold probabilities, were generated by pROC (v1.16.2) in R.

#### Data availability

Sample information and raw sequences analyzed in this study are available in the National Center for Biotechnology Information Sequence Read Archive under accession IDs PRJNA678105, and PRJNA683617 (Table E9). Computer code used in this study is available at https://github.com/BushmanLab/covid_microbiome_2021.

## Supplementary Figures

**Figure E1.**
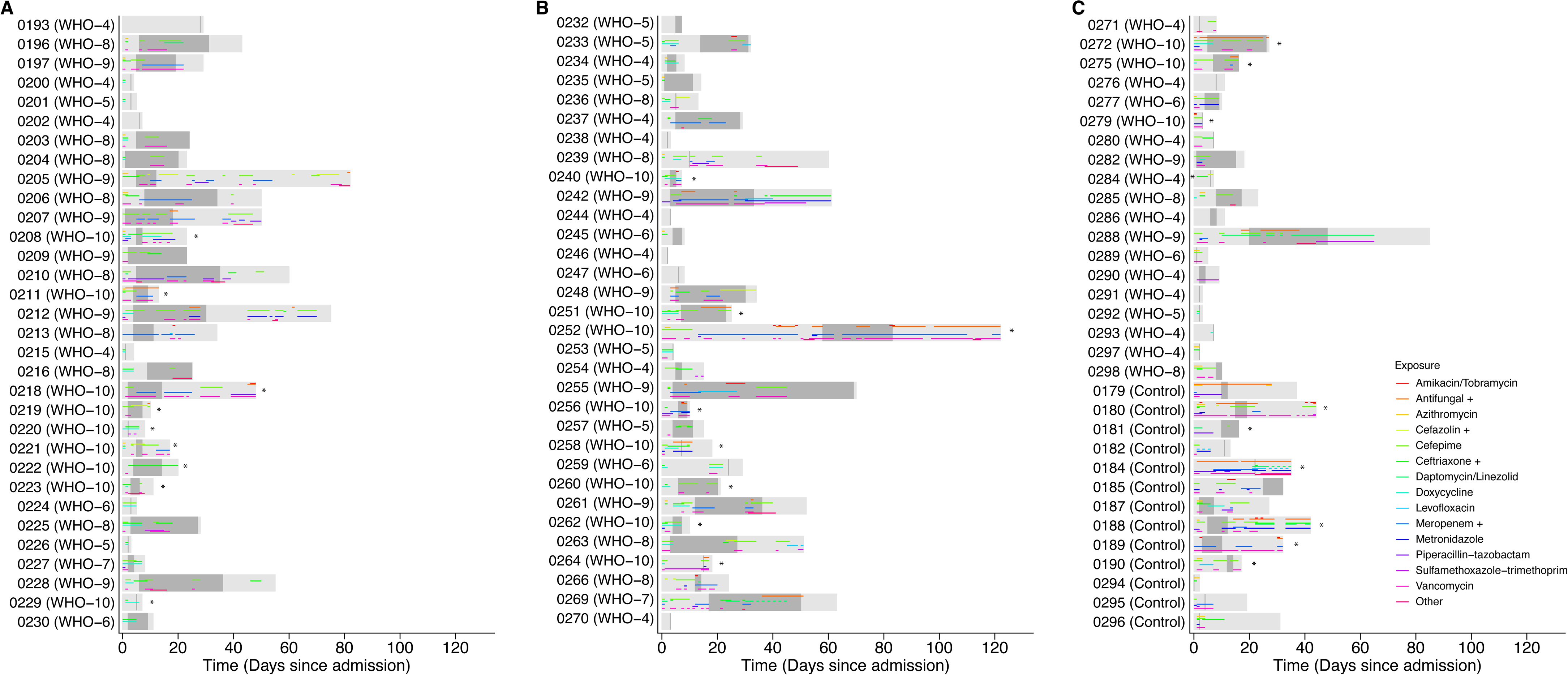
Subject timelines and antibiotic administration. Subjects are grouped by COVID-19 versus non-COVID status. Light gray boxes indicate period of hospitalization and dark gray boxes indicate period of sampling. The X axis indicates time from hospitalization. Maximum COVID-19 disease severity based on WHO score is indicated with subject identifiers, and patients who died (WHO 10) indicated with an asterisk. Antibiotic administration is shown as colored horizontal bars. For simplicity, some antibiotics are grouped with the most common agent within a particular class as indicated by “+”: Cefazolin+ also includes cefalexin and cefadroxil; Ceftriaxone+ also includes ceftazidime and cefpodoxime; Meropenem+ also includes ertapenem and meropenem-vaborbactam. Antifungals include caspofungin, fluconazole, isavuconazonium, posaconazole, voriconazole and atovaquone. “Other” indicates less commonly used antibiotics including amoxicillin, aztreonam, ceftaroline, clindamycin, colistin, fosfomycin, minocycline, amoxicillin-clavulanate, ceftolozane-tazobactum, ampicillin, and ampicillin-sulbactam.

**Figure E2.**
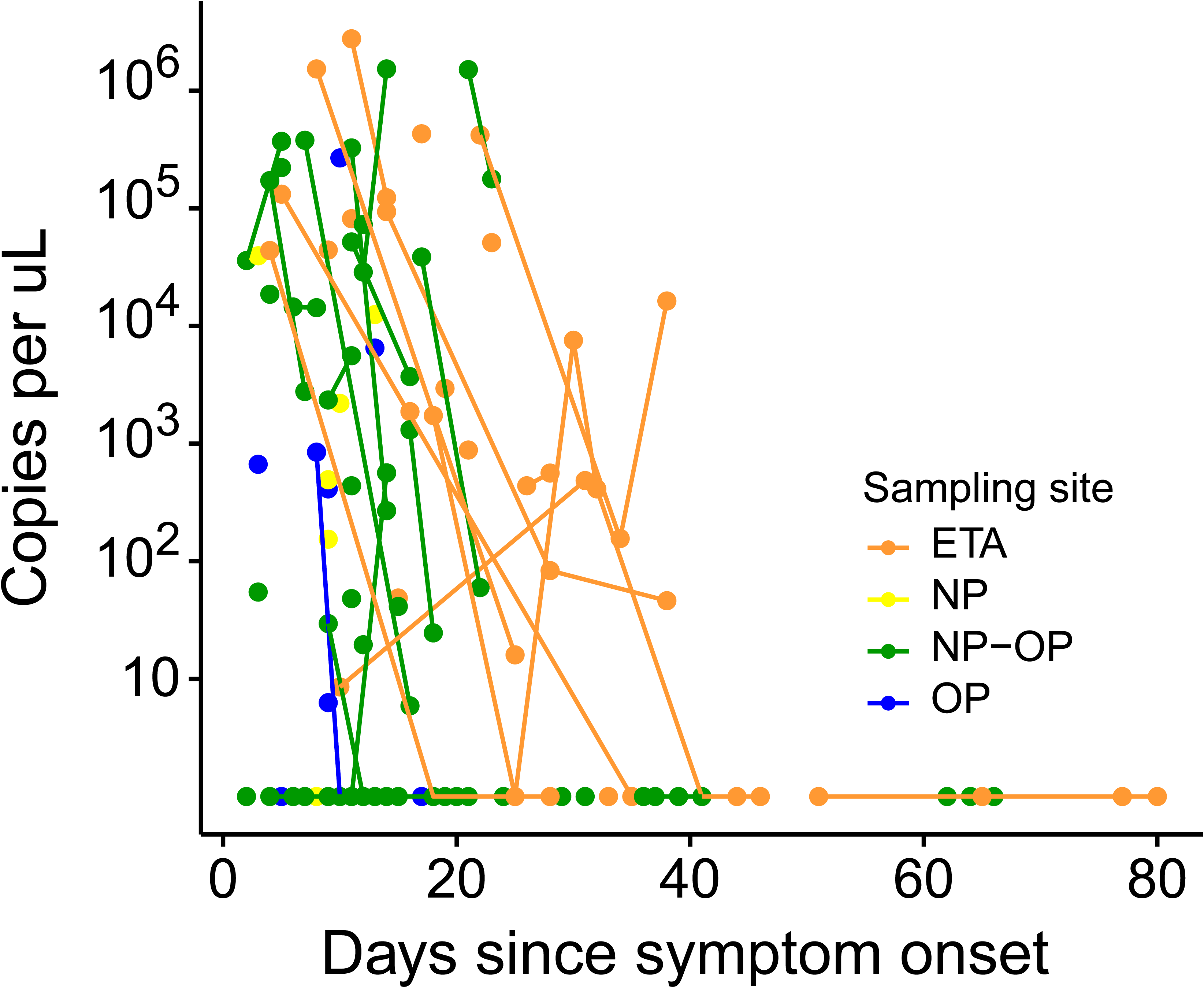
SARS-CoV-2 viral RNA levels in respiratory tract samples. Levels of SARS-CoV-2 were determined by qPCR. Sample type is coded by color, and samples of the same type from the same subject are connected by lines.

**Figure E3.**
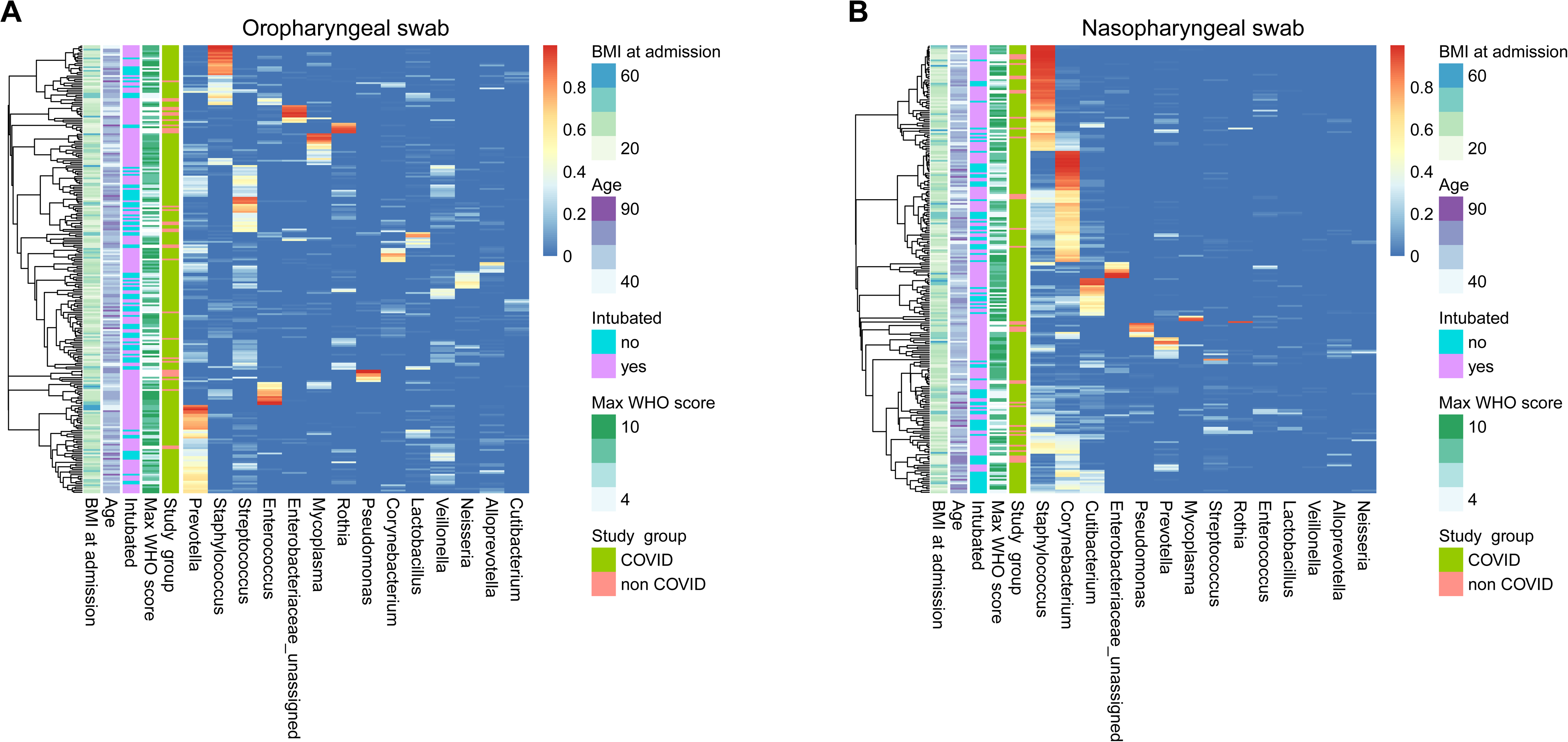
Bacterial communities in oropharyngeal and nasopharyngeal samples. Heatmap showing oropharyngeal **(A)** and nasopharyngeal **(B)** communities.

**Figure E4.**
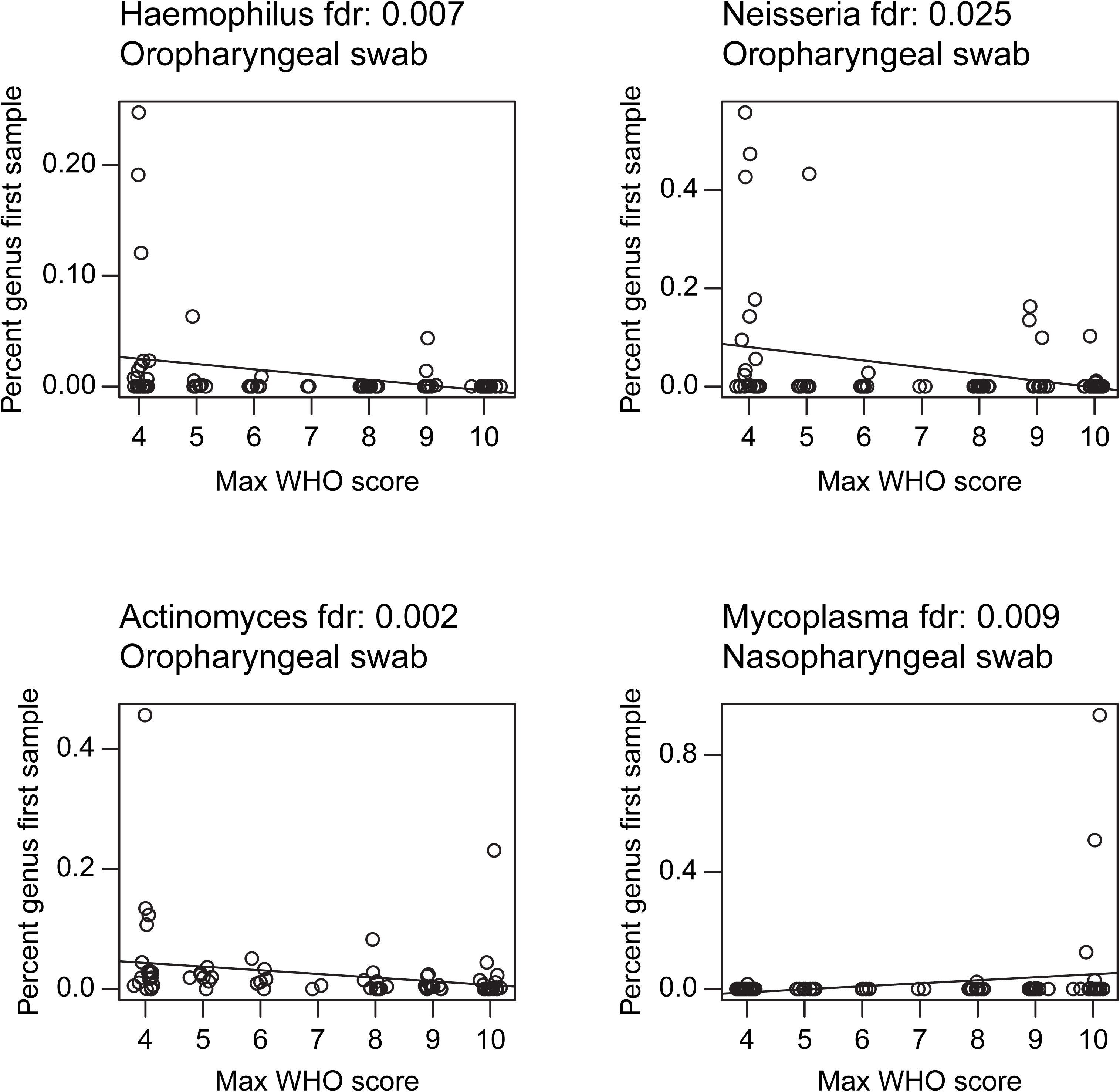
Bacterial taxa present in first sample that are significantly associated with clinical status over course of hospitalization. The x-axis shows the WHO score, the y-axis shows the precent of the community comprised by the indicated genus in the first sample. Sample type and FDR-corrected p-values are shown at the top.

## Supplementary Tables

**Table E1.** Demographic and clinical information on subjects.

**Table E2.** Samples analyzed by 16S rRNA marker gene sequencing.

**Table E3.** Results of qPCR assays to quantify *Anelloviridae* and *Redondoviridae* levels.

**Table E4.** Statistical comparison of bacterial and viral microbiome data to patient demographics, treatment, and outcomes. P values are FDR-corrected and significant associations are highlighted.

**Table E5.** Relationship between Lymphocyte-to-Neutrophil ratios and bacterial and viral microbiome data. To account for multiple daily laboratory tests and day-to-day variability, the first value per calendar day was used, and the average of 3 days (day −1, day 0 and day +1 relative to the microbiome sample) was used. P values are FDR-corrected and significant associations are highlighted.

**Table E6.** Statistical comparison of bacterial and viral microbiome data to clinical laboratory data. P values are FDR-corrected and significant associations are highlighted.

**Table E7**. Statistical comparison of bacterial and viral microbiome data to immune profiling data available on 34 subjects. P values are FDR-corrected.

**Table E8**. Synthetic oligonucleotides used in this study.

**Table E9.** Accession numbers of sequence data generated in this study.

## Notes

**Funding:** This work was supported in part by the Penn Center for Research on Coronaviruses and Other Emerging Pathogens and NIH grant R33-HL137063 (R.G.C., F.D.B.). E.J.W. was supported by NIH grants AI105343, AI082630, the Allen Institute for Immunology and the Parker Institute for Cancer Immunotherapy. J.R.G. was supported by the NIH (T32 CA009140) and a Cancer Research Institute-Mark Foundation Fellowship

### Competing Interest Statement

The authors have declared no competing interest.

### Funding Statement

This work was supported in part by the Penn Center for Research on Coronaviruses and Other Emerging Pathogens and NIH grant R33-HL137063 (R.G.C., F.D.B.). E.J.W. was supported by NIH grants AI105343, AI082630, the Allen Institute for Immunology and the Parker Institute for Cancer Immunotherapy. J.R.G. was supported by the NIH (T32 CA009140) and a Cancer Research Institute-Mark Foundation Fellowship

### Author Declarations

Following informed consent obtained under protocol #823392 approved by the University of Pennsylvania IRB, samples were collected at the Hospital of the University of Pennsylvania

